# Evaluation of Progesterone Resistance and the Role of Downregulation Protocols in Endometrial Preparation for Frozen Embryo Transfer in Patients with Recurrent Implantation Failure—A Retrospective Study

**DOI:** 10.1101/2025.02.22.25322723

**Authors:** Yuxing Luo, Changshuang Cen, Huaicong Liu, Dan Mo, Pingfang Fu, Xin Liu, Lidan Liu, Lansi Wei, Qiuying Gan, Yin Bi, Yihua Yang, Ting Liang

**Author notes:** Ting Liang, Yihua Yang and Yin Bi are co-corresponding authors., **Corresponding author:** Ting Liang, Yihua Yang, Yin Bi. These authors contributed equally to this work (Yuxing Luo, Changshuang Cen). Yuxing Luo and Changshuang Cen are co-first authors. These authors also contributed equally to this work (Yin Bi, Yihua Yang, Ting Liang). **INSTITUTIONAL EMAIL:**.

## Abstract

This study investigates the impact of progesterone resistance on endometrial receptivity and evaluates the efficacy of pituitary downregulation combined with hormone replacement therapy (HRT) in improving pregnancy outcomes following frozen embryo transfer (FET). A retrospective analysis was conducted on 147 patients with recurrent implantation failure (RIF) who underwent secretory-phase endometrial biopsy and subsequent FET treatments at the Reproductive Center of the First Affiliated Hospital of Guangxi Medical University between April 2019 and May 2022. Based on biopsy results and endometrial preparations, patients were categorized into four groups: secretory phase + non-downregulated protocol group (n=29), secretory phase + GnRH-a + HRT Protocol group (n=20), proliferative phase + non-downregulated protocol group (n=59), proliferative Phase + GnRH-a + HRT Protocol group (n=39). Endometrial preparation regimens and pregnancy outcomes during the first FET cycle after biopsy were compared. The results showed that there was no statistical difference in the outcome of assisted pregnancy between the different endometrial preparation protocols in the four groups. These findings suggest that the prioritization of down-regulation protocols may not be essential for FET in patients with RIF, even when endometrial biopsy during the implantation window reveals proliferative-phase characteristics.

## INTRODUCTION

The definition of Recurrent implantation failure (RIF) has not yet been unified. According to Chinese expert consensus, RIF is characterized by the inability to achieve clinical pregnancy following the transfer of at least 3 high-quality embryos in 3 fresh or frozen cycles in adult women under 40 years old [1]. Approximately 10% of patients undergoing IVF treatment will experience RIF [2]. The mechanism of RIF is still unclear, including maternal, male, and embryonic factors, among which maternal immune dysfunction, abnormalities in the anatomy of the reproductive system, chromosomal abnormalities, and endometrial tolerance abnormalities (e.g., displaced receptive window, progesterone resistance) are thought to be the important causes of RIF [3, 4]. The establishment of endometrial receptivity is coordinated primarily by estrogen and progesterone, leading to functional changes in all endometrial cell types through direct and indirect transcriptional and translational regulation [5]. Loss of hormonal homeostasis, disruption of hormone-dependent downstream signaling mechanisms, and/or aberrant inflammation can lead to hormone insensitivity, estrogen dependence/dominance, and progesterone resistance (PR). Progesterone resistance is defined as a low or absent level of progesterone receptor expression that is unable to respond effectively to progesterone stimulation, thus preventing progesterone from working properly, and impeding the differentiation of endometrial stromal cells into decidual cells. Progesterone resistance is considered a crucial factor that impacts endometrial receptivity [6]. Throughout a woman’s menstrual cycle, impaired progesterone signaling and the accumulation of prolonged estrogen action can instigate pathological changes in the endometrium, potentially leading to endometrial-related disorders such as endometriosis and adenomyosis [7, 8].In this study, we proposed to investigate the effect of progesterone resistance on the establishment of endometrial receptivity and to assess the effect of pituitary downregulation combined with hormone replacement therapy (HRT) on the outcome of assisted pregnancy in patients with RIF by conducting a retrospective clinical analysis of RIF patients.

## MATERIALS AND METHODS

### Study design and population

Clinical data were collected from patients with RIF who underwent endometrial biopsy in the secretory phase and received IVF/ICSI-ET or FET treatment at the Reproductive Center of the First Affiliated Hospital of Guangxi Medical University and Guigang People’s Hospital, between April 2019 and May 2022. The flow of this study is shown in **FIGURE 1 (FLOW CHART OF THIS STUDY)**. The study was approved by the Ethics Committee of the First Affiliated Hospital of Guangxi Medical University and Ethics Committee of the Guigang People’s Hospital (approval number: 2024-E0850, ELW-2024-029-01).

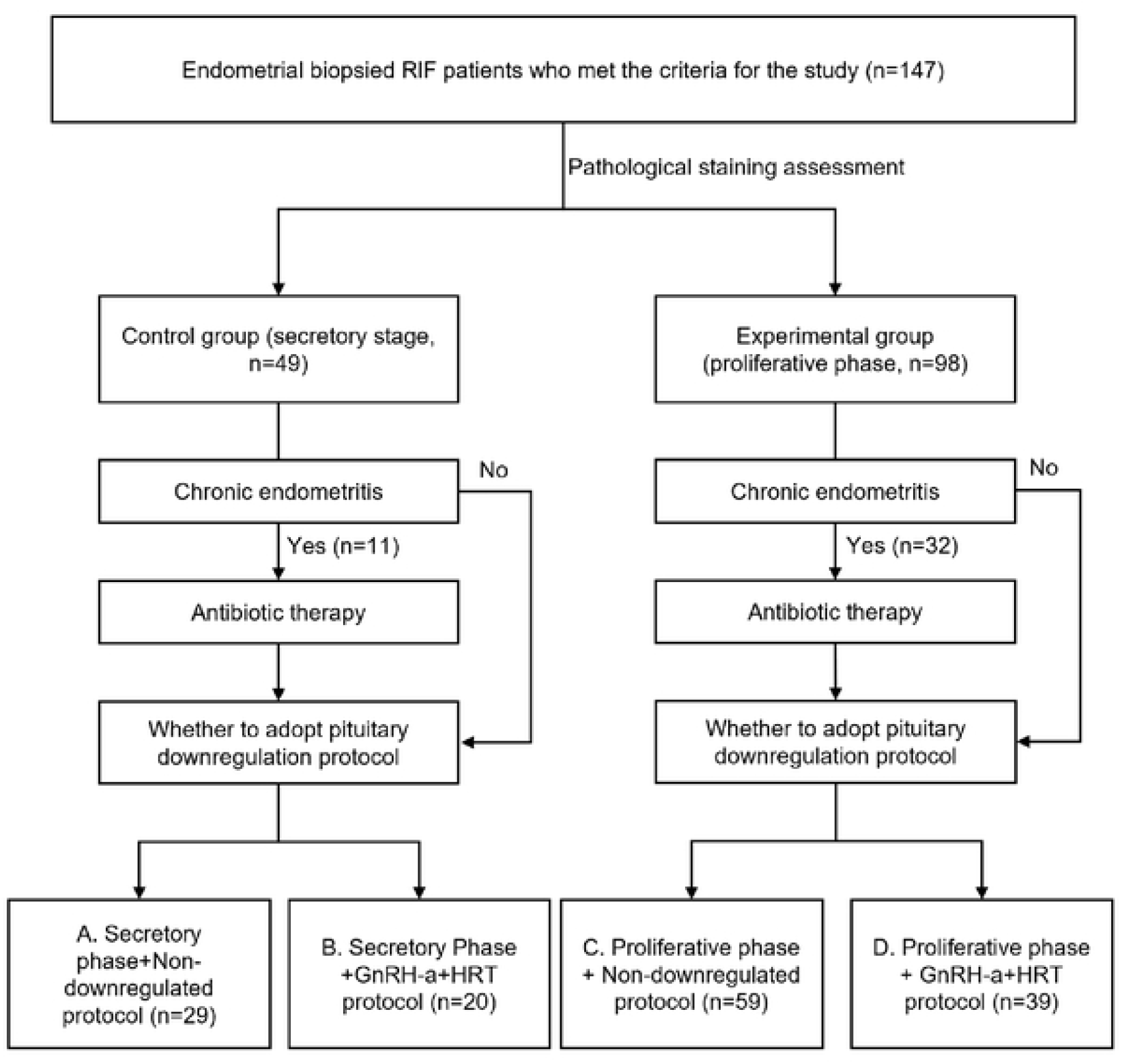

Patients included in the study were required to meet the following criteria: (1) Women aged 20 to 40; (2) A clinical pregnancy could not be achieved after transferring at least three high-quality embryos in three fresh or frozen cycles, and high-quality embryos were defined as day 3 embryos (≥ 8 cells, uniform cleavage stage, and <10% fragmentation) and blastocysts (≥ 3BB); (3) progesterone level test on the day of endometrial biopsy suggests the presence of ovulation in that cycle; (4) the first FET cycle after endometrial biopsy.

Patients with the following conditions were excluded: (1)Presence of imaging-confirmed adenomyosis, polycystic ovary syndrome (PCOS), aand other disorders in which progesterone resistance is clearly present; (2) combined with a confirmed diagnosis of thyroid dysfunction, Cushing’s syndrome, congenital adrenocortical hyperplasia, and other diseases that may affect the secretion of steroid hormones; (3) during the same period of time, the application of other medications that could potentially affect the secretion of steroid hormones; (4) abnormalities of the uterine cavity morphology, including uterine abnormalities (longitudinal uterus, unicornuate uterus, double uterus), submucosal uterine fibroids or interstitial fibroids larger than 5cm, or uterine adhesions, etc.; (5) the presence of contraindications to Assisted Reproductive Technology (ART) and pregnancy, or conditions that severely affect pregnancy outcomes, such as uterine fibroids; (6) insufficient clinical data or inability to carry out long-term follow-ups.

### Study methods

Whether the endometrial biopsy was taken in the secretory phase was initially determined by the last menstruation and menstrual cycle, and the day of ovulation was determined by daily urine LH tests starting from the 12th day of menstruation, with a strong positive LH for D0, and endometrial biopsy was performed on D7 with the simultaneous measurement of the progesterone level (>10 ng/ml). After endometrial biopsy, if CD138-positive cells ≥5 HPF, it suggests the presence of chronic endometritis, and two cycles of antibiotics (doxycycline + metronidazole) are required before FET [9]. If chronic endometritis is not present, direct FET may be considered. We then compared the endometrial preparation protocols and pregnancy outcomes in the first FET cycle after endometrial biopsy.

### Endometrial preparation protocols

The endometrial preparation protocols for FET were categorized into non-downregulation and pituitary downregulation groups. Non-downregulation groups include the hormone replacement (HRT) protocol, the natural cycle (NC) protocol, and the ovulation induction cycle (OI) protocol.

#### (1) HRT protocol

On the third day of menstruation, estrogen therapy was initiated and continued until the endometrial thickness reached ≥ 7 mm. Subsequently, progesterone was administered intramuscularly to induce endometrial transformation.

#### (2) NC protocol

When the follicle diameter was ≥ 18 mm, and the endometrial thickness was ≥ 7 mm, the ovulation was induced by intramuscular injection of chorionic gonadotropic hormone (HCG) at 10000U as necessary.

#### (3) OI protocol

Letrozole tablets 2.5-5 mg/d were administered orally for 5 consecutive days on the 3rd to 5th day of menstruation. When the follicle diameter was ≥18 mm and the endometrial thickness was ≥7 mm, ovulation was induced by intramuscular injection of HCG 10000U. Ovulation was monitored by ultrasound two days later, and if it was ovulated, progesterone was given to transform the endometrium by intramuscular injection.

#### (4) GnRH-a+HRT protocol

On the 2nd to 3rd day of the menstrual cycle, a subcutaneous injection of 3.75 mg long-acting triptorelin acetate was administered. After 28 to 30 days, a follow-up was conducted, including vaginal ultrasound and hormone level measurements. If the ultrasound indicated an endometrial thickness of <5mm, no follicles with a diameter >10mm, and no cysts, and the hormone levels showed FSH<5U/L, LH<5U/L, and E2<50pg/L, then the GnRH-a + HRT protocol was adopted for endometrial preparation and embryo transfer.

### Patient groupings

In this study, patients were divided into four groups based on endometrial preparation protocols and menstrual cycle phase:

#### (1) Secretory Phase + Non-Downregulated Protocol (n=29)

This group consisted of patients in the secretory phase who received a protocol without downregulation.

#### (2) Secretory Phase + GnRH-a + HRT Protocol (n=20)

This group consisted of patients in the secretory phase who received GnRH-a+HRT protocol.

#### (3) Proliferative Phase + Non-Downregulated Protocol (n=59)

This group included patients in the proliferative phase who were treated with a protocol without downregulation.

#### (4) Proliferative Phase + GnRH-a + HRT Protocol (n=39)

This group comprised patients in the proliferative phase who received GnRH-a+HRT protocol.

### Definition of pregnancy outcome

#### (1) Biochemical pregnancy

peripheral β-HCG was measured 2 weeks after embryo transfer (>5.8 U/L was considered positive) but no gestational sac was seen on ultrasound later on.

#### (2) Clinical pregnancy

transvaginal ultrasound performed 4-5 weeks after embryo transfer to see the gestational sac, including intrauterine pregnancy, ectopic pregnancy, and simultaneous intrauterine and ectopic pregnancy.

#### (3) Ectopic pregnancy

transvaginal ultrasound performed 4-5 weeks after embryo transfer shows that the gestational sac was deposited outside the uterine cavity.

#### (4) Miscarriage

loss of pregnancy at less than 28 weeks of gestation when the fetus weighs less than 1000g.

#### (5) Live birth

at least one newborn born alive at 28 weeks of gestation.

### Statistical analysis

Statistical software SPSS 26.0 was used to analyze the data. Normally distributed continuous variables were expressed as mean ± standard deviation (x ± s), and differences between four groups were compared using one-way analysis of variance (ANOVA) assess whether there were statistically significant differences among the group means. Post-hoc tests (e.g., Tukey’s HSD) were conducted following a significant ANOVA result to identify specific group differences. Variables with skewed distributions were expressed as median ± interquartile spacing, and differences between groups were tested by nonparametric methods. The difference in results was statistically significant as P < 0.05.

## RESULTS

### General baseline information of patients

A total of 147 patients with RIF met the study criteria. According to the data presented in TABLE 1, we can see that there was no statistical difference in age (33.45±3.460 vs. 33.60±7.943 vs. 32.97±5.5153 vs. 32.97±5.515, p=0.956), BMI (22.19±2.552 vs. 21.77±2.944 vs. 22.04± 2.587 vs. 21.35±2.765, p=0.545), and duration of infertility (4.17±3.263 vs. 5.95±3.706 vs. 4.76 ± 2.932 vs. 5.23 ± 4.428, p=0.340) in the control and experimental groups, indicating that the follow-up indicators were comparable.

**TABLE 1.**
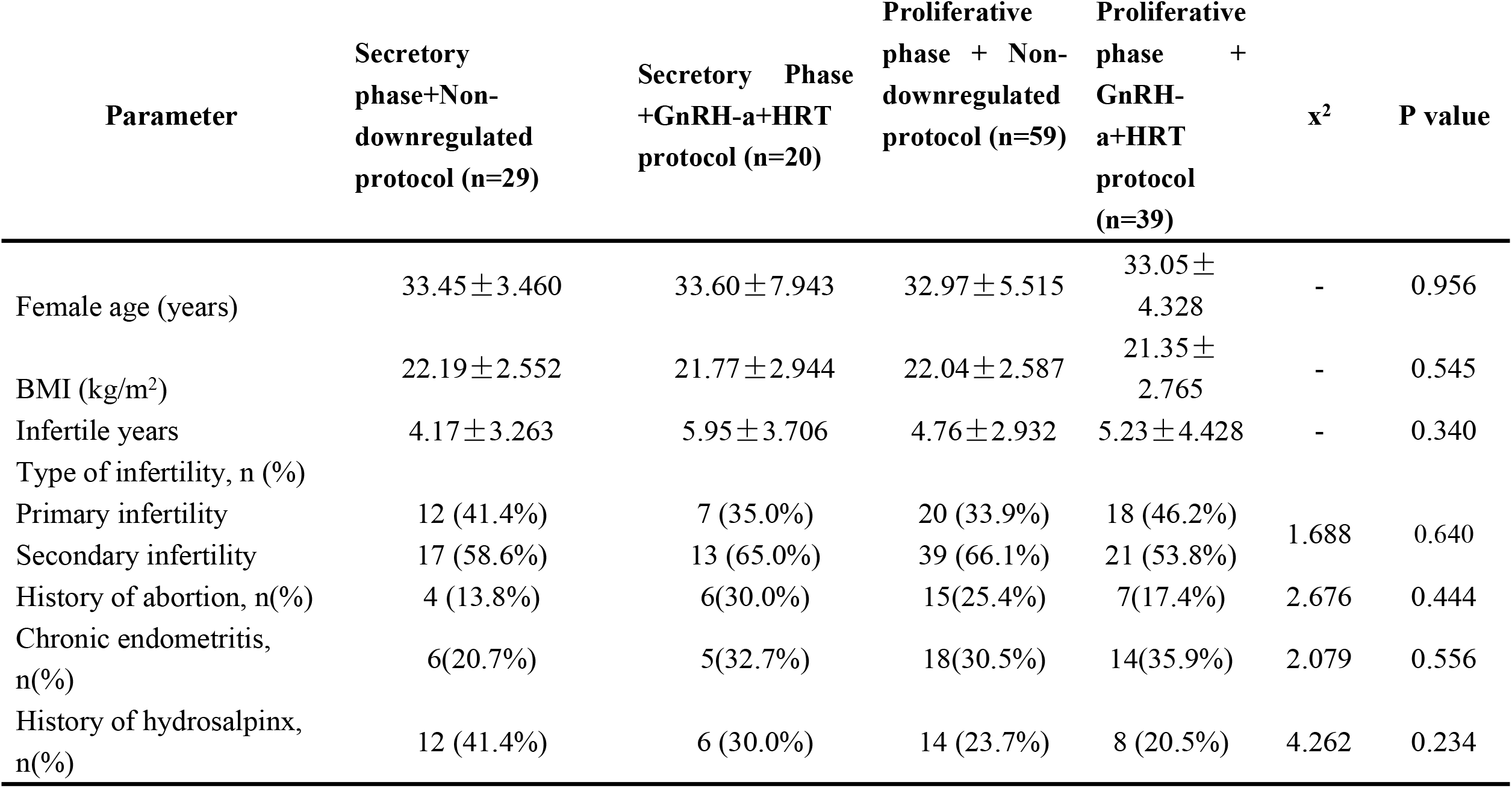
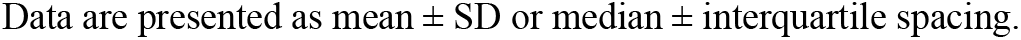
GENERAL BASELINE INFORMATION OF PATIENTS.

| Parameter | Secretory phase+Non-downregulated protocol (n=29) | Secretory Phase +GnRH-a+HRT protocol (n=20) | Proliferative phase + Non-downregulated protocol (n=59) | Proliferative phase + GnRH-a+HRT protocol (n=39) | $\chi^2$ | P value |
| --- | --- | --- | --- | --- | --- | --- |
| Female age (years) | $33.45 \pm 3.460$ | $33.60 \pm 7.943$ | $32.97 \pm 5.515$ | $33.05 \pm 4.328$ | - | 0.956 |
| BMI (kg/m<sup>2</sup>) | $22.19 \pm 2.552$ | $21.77 \pm 2.944$ | $22.04 \pm 2.587$ | $21.35 \pm 2.765$ | - | 0.545 |
| Infertile years | $4.17 \pm 3.263$ | $5.95 \pm 3.706$ | $4.76 \pm 2.932$ | $5.23 \pm 4.428$ | - | 0.340 |
| Type of infertility, n (%) |  |  |  |  |  |  |
| Primary infertility | 12 (41.4%) | 7 (35.0%) | 20 (33.9%) | 18 (46.2%) | 1.688 | 0.640 |
| Secondary infertility | 17 (58.6%) | 13 (65.0%) | 39 (66.1%) | 21 (53.8%) |  |  |
| History of abortion, n(%) | 4 (13.8%) | 6(30.0%) | 15(25.4%) | 7(17.4%) | 2.676 | 0.444 |
| Chronic endometritis, n(%) | 6(20.7%) | 5(32.7%) | 18(30.5%) | 14(35.9%) | 2.079 | 0.556 |
| History of hydrosalpinx, n(%) | 12 (41.4%) | 6 (30.0%) | 14 (23.7%) | 8 (20.5%) | 4.262 | 0.234 |

Data are presented as mean ± SD or median ± interquartile spacing.

### First FET preparation after endometrial biopsy in RIF patients

The basal steroid hormone levels (bFSH, bLH, bE_2_) and AMH levels of the patients in the experimental group were comparable with those of the control group. For endometrial preparation, there was no statistically significant difference in endometrial thickness on the day of conversion between the four groups (9.10±0.92 vs. 9.55±1.45 vs. 8.95±1.43 vs. 9.17±1.29, p=0.242). But progesterone on the day of biopsy were significantly lower than those in the secretory phase in both groups of patients whose biopsies were in the proliferative phase, regardless of whether or not down-regulation treatments were performed during endometrial preparation (14.86±8.00 vs. 13.13±4.40 vs. 6.51±5.47 vs. 4.28±3.11, p<0.001). There was no statistically significant difference in the quality of transferred embryos between the four groups for either cleavage stage embryos or blastocysts (TABLE 2).

**TABLE 2.** FIRST FET ASSISTED PREGNANCY AFTER ENDOMETRIAL BIOPSY IN FOUR GROUPS OF PATIENTS.

| Parameter | Secretory<br>phase+Non-<br>downregulated<br>protocol (n=29) | S+GnRH-<br>a+HRT<br>protocol<br>(n=20) | Proliferative<br>phase + Non-downregulated<br>protocol (n=59) | Proliferative<br>phase + GnRH-a+HRT protocol<br>(n=39) | x<sup>2</sup> | P<br>value |
| --- | --- | --- | --- | --- | --- | --- |
| bFSH (U/L) | $6.40 \pm 1.91$ | $6.31 \pm 2.66$ | $6.74 \pm 2.42$ | $5.28 \pm 1.85$ | - | 0.018 |
| bLH (U/L) | $7.25 \pm 6.16$ | $6.10 \pm 4.06$ | $8.33 \pm 7.03$ | $5.71 \pm 3.73$ | - | 0.142 |
| bE<sub>2</sub> (pg/ml) | $96.04 \pm 92.53$ | $101.98 \pm 100.47$ | $154.62 \pm 451.82 \pm$ | $120.72 \pm 104.09$ | - | 0.806 |
| AMH (ng/ml) | $2.60 \pm 1.28$ | $7.78 \pm 21.08$ | $3.93 \pm 3.02$ | $3.38 \pm 1.80$ | - | 0.139 |
| Endometrial thickness on<br>transformation day (mm) | $9.10 \pm 0.92$ | $9.55 \pm 1.45$ | $8.95 \pm 1.43$ | $9.17 \pm 1.29$ | - | 0.242 |
| Progesterone on biopsy<br>day (ng/ml) | $14.86 \pm 8.00$ | $13.13 \pm 4.40$ | $6.51 \pm 5.47$ | $4.28 \pm 3.11$ | - | 0.000 |
| Type of embryos<br>transferred, n (%) |  |  |  |  |  |  |
| D3 cleavage-stage<br>embryos, n (%) | 19 (65.5%) | 15 (75.0%) | 33 (55.9%) | 21 (53.8%) | 3.260 | 0.353 |
| Blastocyst, n (%) | 10 (34.5%) | 5 (25.0%) | 26(44.1%) | 18(46.2%) |  | 0.187 |
| Number of ET embryos | $1.66 \pm 0.484$ | $1.70 \pm 0.470$ | $1.73 \pm 0.478$ | $1.67 \pm 0.462$ | - | 0.881 |
| Pregnancy outcomes, n<br>(%) |  |  |  |  |  |  |
| Biochemical pregnancy | 4 (13.8%) | 3 (15.0%) | 6 (10.2%) | 6 (15.4%) | 0.704 | 0.872 |
| Clinical pregnancy | 7 (24.1%) | 5 (25.0%) | 24 (40.7%) | 15 (38.5%) | 3.433 | 0.330 |
| Ectopic pregnancy | 1 (3.4%) | 1 (5.0%) | 1 (1.7%) | 0 (0.0%) | 2.011 | 0.570 |
| Miscarriage | 0 (0.0%) | 1 (20.0%) | 1 (4.2%) | 1 (12.5%) | 2.720 | 0.437 |
| Live birth | 7 (24.1%) | 4 (20.0%) | 23 (39.0%) | 13 (33.3%) | 3.503 | 0.320 |

Data are presented as mean ± SD or median ± interquartile spacing.

### Effect of a pituitary downregulation protocol on FET outcomes in patients with RIF

As can be seen in the table, the biochemical pregnancy rate (13.8% vs. 15.0% vs. 10.2% vs. 15.4%, p=0.872), clinical pregnancy rate (24.1% vs. 25.0% vs. 40.7% vs. 38.5%, p=0.330), ectopic pregnancy rate (3.4% vs. 5.0% vs. 1.7% vs. 0.0%, p=570), miscarriage rate (0.0% vs. 20.0% vs. 4.2% vs. 12.5%, p=0.437), and live birth rate in four groups (24.1% vs. 20.0% vs. 39.0% vs. 33.3%, p=0.320) were not statistically different (TABLE 2).

## DISCUSSION

With the rapid development of assisted reproductive techniques and the application of PGT technology, significant progress has been made in the acquisition and screening of high-quality embryos. However, the embryo implantation and live-birth rates of in vitro fertilization-embryo transfer (IVF-ET) remain relatively low. It is estimated that up to two-thirds of all implantation failures are due to defects in endometrial receptivity [10]. Endometrial receptivity refers to the ability of the endometrium to accept an embryo implantation within a specific time window, during which the trophoblast ectoderm of the blastocyst can attach to endometrial epithelial cells and subsequently invade the endometrial mesenchyme and vascular system [5]. The establishment of endometrial receptivity requires the coordinated action of estrogen and progesterone, and progesterone resistance is caused by progesterone receptor defects, receptor activator alterations, related genetic changes, etc., which reduce or deprive patients of progesterone sensitivity [11]. Studies have shown that progesterone resistance may affect endometrial receptivity, which in turn affects embryo implantation, leading to female infertility [12].

Progesterone resistance is commonly observed in conditions such as endometriosis, PCOS, and endometrial hyperplasia. The causal relationships among these conditions are not fully elucidated at present. In these diseases, inflammatory factors, microRNAs (miRNAs), DNA methylation, leukemia inhibitory factor (LIF), and Homeobox gene A10 (HOXA10) are involved in the regulation of PR, which further mediates the development of progesterone resistance [12, 13]. In endometriosis, signaling pathways associated with proliferation and cell survival are activated, while the anti-proliferation progesterone pathway is shut down. Progesterone resistance leads to insufficient antagonism of estrogen action, increased inflammation, insufficient mesenchymal differentiation, and endometrial remodeling, all of which can result in the endometrium being unreceptive for embryo implantation. [14]. Hydrosalpinx is the result of chronic inflammation of the fallopian tube leading to adhesion and atresia of the umbilical end of the tube and accumulation of inflammatory exudate. Under normal physiological conditions, estrogen promotes the development of the tubal musculature and the secretion of epithelium, and enhances the amplitude of rhythmic contractions of the tubal muscle, while progesterone has the opposite effect. During the IVF superovulation process, the amount of fluid accumulation may increase significantly as estrogen levels continue to rise. The low pregnancy rate associated with hydrosalpinx is mainly due to impaired embryo implantation, embryotoxicity, mechanical washout, and alterations in the intrauterine environment [15]. Hydrosalpinx causes damage not only to functional endometrial cells but also to basal cells, leading to decreased expression of HOXA10 in proliferating endometrial and decidual cells, which in turn impairs endometrial receptivity and leads to impaired implantation of embryos [16]. In our study, there was no statistically significant difference in the incidence of hydrosalpinx in four groups, probably because the number of cases included was too small.

Frozen-thawed embryo transfer is an important part of assisted reproductive technology, and choosing the proper endometrium preparation protocol is crucial. At present, NC protocol and HRT protocol are the two most commonly employed methods for endometrial preparation in FET cycles. The NC protocol is mainly used for patients with regular ovulation. The HRT protocol is mostly used for patients with ovulation disorders, thin endometrium, and follicular development disorder. However, this protocol cannot guarantee complete pituitary suppression, and thus, dominant follicle development may occur during the administration of the medication, which may cause an early onset of LH peaks, interfering with the internal environment for embryo implantation [17, 18]. The addition of GnRH-a downregulation to HRT cycles minimizes early onset LH peaks. The results of this study showed that there was no statistical difference in the outcome of assisted pregnancy between the different endometrial preparation protocols in the four groups. This result suggests that, for patients with impaired endometrial transformation, the use of a down-regulation regimen during endometrial preparation might has no significant impact on subsequent FET pregnancy outcomes.

In summary, RIF is a challenge for assisted reproduction technology. Existing studies have shown that downregulation of progesterone receptor expression in the endometrium can reduce or eliminate the sensitivity of progesterone, leading to changes in endometrial receptivity and a decrease in the rate of implantation of embryos, ultimately resulting in infertility [6]. Therefore, exploring the mechanism of endometrial receptivity formation, analyzing its related influencing factors, and studying the interactions among the factors are still the focus of the current research work. The GnRH-a+HRT protocol is a newer endometrial preparation way compared to natural cycles and HRT cycles, which is especially suitable for infertile patients with PCOS, adenomyosis, RIF, and chronic endometritis. However, the prioritization of down-regulation protocols may not be essential for FET in patients with RIF. This study is a single-center retrospective case study with a small sample size, which requires large multi-center clinical data and basic experiments for further exploration.

## CONCLUSION

The prioritization of down-regulation protocols may not be essential for FET in patients with RIF, even when endometrial biopsy during the implantation window reveals proliferative-phase characteristics.

## Data Availability

Data cannot be shared publicly because of the protection of patient data privacy. Data are available from the Ethics Committee of the First Affiliated Hospital of Guangxi Medical University and Ethics Committee of the Guigang People's Hospital for researchers who meet the criteria for access to confidential data.

## ACKNOWLEDGEMENTS

We would like to thank the patients and staff of the Reproductive Medicine Center, the First Affiliated Hospital of Guangxi Medical University for their contribution.

## AUTHOR CONTRIBUTION STATEMENT

Yuxing Luo was responsible for the data curation, formal analysis and writing original draft preparation. Changshuang Cen was responsible for data collection, statistical analysis, and manuscript revision. Yuxing Luo and Changshuang Cen contributed equally to this work and share first authorship. Huaicong Liu, Dan Mo and Pingfang Fu contributed to the revision of the paper. Xin Liu, Lidan Liu, Lansi Wei, and Qiuying Gan contributed to data collection. Yin Bi critically revised the manuscript for important intellectual content. Yihua Yang conceptualized the entire research framework and provided financial support. Ting Liang provided guidance in data analysis, revised the manuscript’s content and funding acquisition.

## Notes

### Competing Interest Statement

The authors have declared no competing interest.

### Funding Statement

Yes

### Author Declarations

The study was approved by the Ethics All the authurs ensured that we had discussed whether all data were fully anonymized before we accessed them. The need for consent was waived by the ethics committee and the information is included. Committee of the First Affiliated Hospital of Guangxi Medical University and Ethics Committee of the Guigang People's Hospital (approval number: 2024-E0850, ELW-2024-029-01). All authors had not access to information that could identify individual participants during or after data collection.

